# Investigation of the protection efficacy of face shields against aerosol cough droplets

**DOI:** 10.1101/2020.07.06.20147090

**Authors:** A. Ronen, H. Rotter, S. Elisha, S. Sevilia, B. Parizer, N. Hafif, A. Manor

## Abstract

Simple plastic face shields have numerous practical advantages over regular surgical masks. In light of the spreading COVID-19 pandemic, the potential of face shields as a substitution for surgical masks, as a recommendation to the general population, was investigated In order to determine the efficacy of the protective equipment we used a cough simulator that was carefully tuned to replicate human cough in terms of droplet size distribution and jet velocity. The protective equipment considered was placed on a manikin head that simulated human breathing. An Aerodynamic Particle Sizer (APS) was used to analyze the concentration and size distribution of small particles that reached the manikin respiration pathways. Additionally, water sensitive papers were taped on the tested protective equipment and the manikin face, and subsequently photographed and analyzed. In the case of frontal exposure, for droplet diameter larger than 3 μm, the shield efficiency in blocking cough droplets was found to be comparable to that of regular surgical masks, with enhanced protection for portions of the face that the mask does not cover. Additionally, for finer particles, down to 0.3 micron diameter, a shield was found to perform even better, blocking about 10 times more fine particles than the surgical mask. When exposure from the side was considered, the performance of the shield was found to depend dramatically on its geometry. While a narrow shield allowed more droplets and aerosol to penetrate in comparison to a mask under the same configuration, a slightly wider shield significantly improved the performance. The ability of a shield worn by an infected person in order to protect others in his vicinity was also investigated. A shield, and alternatively, a surgical mask, were placed on the cough simulator, while the breathing simulator remained totally exposed. In both cases, no droplets or particles were found in the vicinity of the breathing simulator.

## INTRODUCTION

Pathogen-laden aerosol transport triggered by expiratory events such as coughing, sneezing, or talking, fundamentally contributes to the spread of the disease and to its potential of becoming an epidemic (Jones. 2020, Morawska and Milton. 2020). The study of the processes involved in such expiratory events and investigation of the efficiency of related personal protective equipment (PPE) are therefore crucial.

The COVID-19 pandemic that has spread worldwide in just a few months has challenged public health policymakers in many different aspects. The ultimate objective for controlling the pandemic on the national level is a lowering of the infection rate, such that on average, an infected person will infect less than one other person, and thus eventually end the spread of the disease. In this respect, lowering the rate of spread, even modestly, can greatly contribute to the overall control of the disease. For that reason, PPE that is considered insufficient in the occupational context, focusing on the well-being of healthcare workers operating in a highly contaminated environment, can make a considerable difference when the problem at hand is controlling an epidemic over mass-population scales.

The advantages of face shields over regular surgical masks are numerous. While surgical masks have limited availability and are disposable, face shields can be reused and are easily cleaned. They are comfortable to wear, retain less dermal facial heat, have no impact on breathing resistance, are less claustrophobic and inexpensive (Roberge. 2016). They reduce the potential for autoinoculation by preventing the wearer from touching their face, and essentially protect the entire face and not only the expiratory pathway (CDC and NIOSH. 2015). Also, use by a large fraction of the population of protective equipment which is slightly less effective but more comfortable or simple to wear may be more beneficial, in the public health context, than more effective equipment which is less comfortable or cumbersome. With these considerations, a face shield, although only partially protecting the users, can reduce the rate of infection, and may be adopted by more users due to its relative comfort (Perencevich et al. 2020).

Motivated by these concepts, this work examines the potential of face shields as the sole protective equipment used by the entire population, instead of surgical masks. It is important to emphasize that for health-care workers face shields should generally not be used alone, but rather in conjunction with other protective equipment and are therefore classified as adjunctive PPE (Roberge, 2016).

Droplet formation during an expiratory event like coughing, sneezing, talking, or even breathing, is a complicated process which involves complex flow through the different expiratory pathways. When a rapid airflow passes over a wet boundary, strips of liquid are lifted from the boundary, and then shredded and torn into droplets. The final droplet size distribution depends upon the fluid velocity, the pathways’ topology, the boundary wetting, and physical attributes of the mucus (Gralton, et al., 2011). Spray created by different expiratory events has been studied in different aspects. Yang, et al., (2007) measured the cough droplets size distribution by having volunteers cough into airbags, and subsequently analyze the droplet size of the contents. Their measurements yield a droplet number size distribution with a mode at 8.35 µm. More controlled experiments conducted by Morawska, et al. (2009) took into account the droplet dynamics between exhalation and measurements and found a different size distribution. The size distributions observed for different expiratory events were measured, and the authors suggest that every observed distribution is a combination of four distinct distributions, each characterized by a specific mode. Specifically, a cough event involves two main modes, at 0.8 µm and at 1.8 µm.

The ejected air stream during a cough event creates a jet. The velocity immediately outside the mouth is greatest, and decreases further downstream. Specifying a jet velocity is therefore a matter of definition. The common way of defining and measuring the velocity in coughing and sneezing events is to estimate arrival times at a given distance using imagery techniques. Tang, et al., (2013) used a shadowgraph imaging technique combined with high speed photography and reported velocities around 4.5 m/s for coughs.

An earlier comprehensive work which studied the level of protection provided by face shields (Lindsley, et al., 2014; 2010; 2013) used a cough simulator, a breathing simulator, and an optical particle counter to estimate the amount of spray inhaled by a susceptible person in the vicinity of a cough event. The inhaled, droplet-laden air was transferred into the particle analyzer, and the total inhaled mass was determined as a function of time. The study concludes that wearing a face shield substantially reduces the number of inhaled particles in the short term, but in the longer term, very small particles bypass the shield and are inhaled.

The current study utilizes a cough simulator and besides monitoring inhaled fine particles, also provides further insight into several aspects. First, infected droplets can impact on other parts of the face and initiate subsequent transmittance. To address this, water-sensitive papers (see below) were taped on the manikin head and on the tested PPE. This method enables the simultaneous assessment of the spatial distribution of droplets impacting different parts of the face. Second, several configurations of the simulators were tested, including height and orientation differences. Finally, a shield was placed on the coughing simulator itself to assess the amount of protection provided to adjacent individuals when an infected person is wearing the shield. This issue is very important in the current context since COVID-19 can be transmitted by non-symptomatic infected individuals.

It is important to stress that there are significant differences in all parameters of the ejected spray formed by expiratory events among different individuals and different scenarios. Therefore, the typical parameters chosen for this work do not represent every individual case.

It is also worth noting that studying the fate of droplets and aerosol originating in an expiratory event is by itself insufficient to assess probability of infection. Droplets of different sizes are created in different locations in the respiratory tract and, consequently, pathogen load is non-uniformly distributed among different droplet sizes in a way that depends on the specific pathogen and expiratory activity (Gralton et al. 2011, Fennelly. 2020). Therefore, this work focuses on the comparison between different PPE, rather than in assessing the probability of transmittance.

## METHODS

The experiment was performed in a 200 m^3^, air conditioned room, with the experimental setup (see Figures 1 and 2) positioned in the middle of the room. Typical background flow velocity in the room is 2-8 cm/sec. Background particle number concentrations were in the range of 2-4 particles/cm^3^. An airbrush diffuser [Iwata Eclipse SBS] was used as a source of droplets spray. Controlling the operation duration (1 second) allowed calibration of the amount of released liquid. The pressure of the diffuser (here, 3 bars) was adjusted to provide a jet speed of about 5 m/s, typical for a cough event (Tang, et al., 2013). The ratio between the flow rate of air and liquid into the instrument was calibrated to obtain the desired size distribution. The released mass in each of the cough simulations was 100 μl, which is the upper limit expected from a real cough.

**Figure 1.**
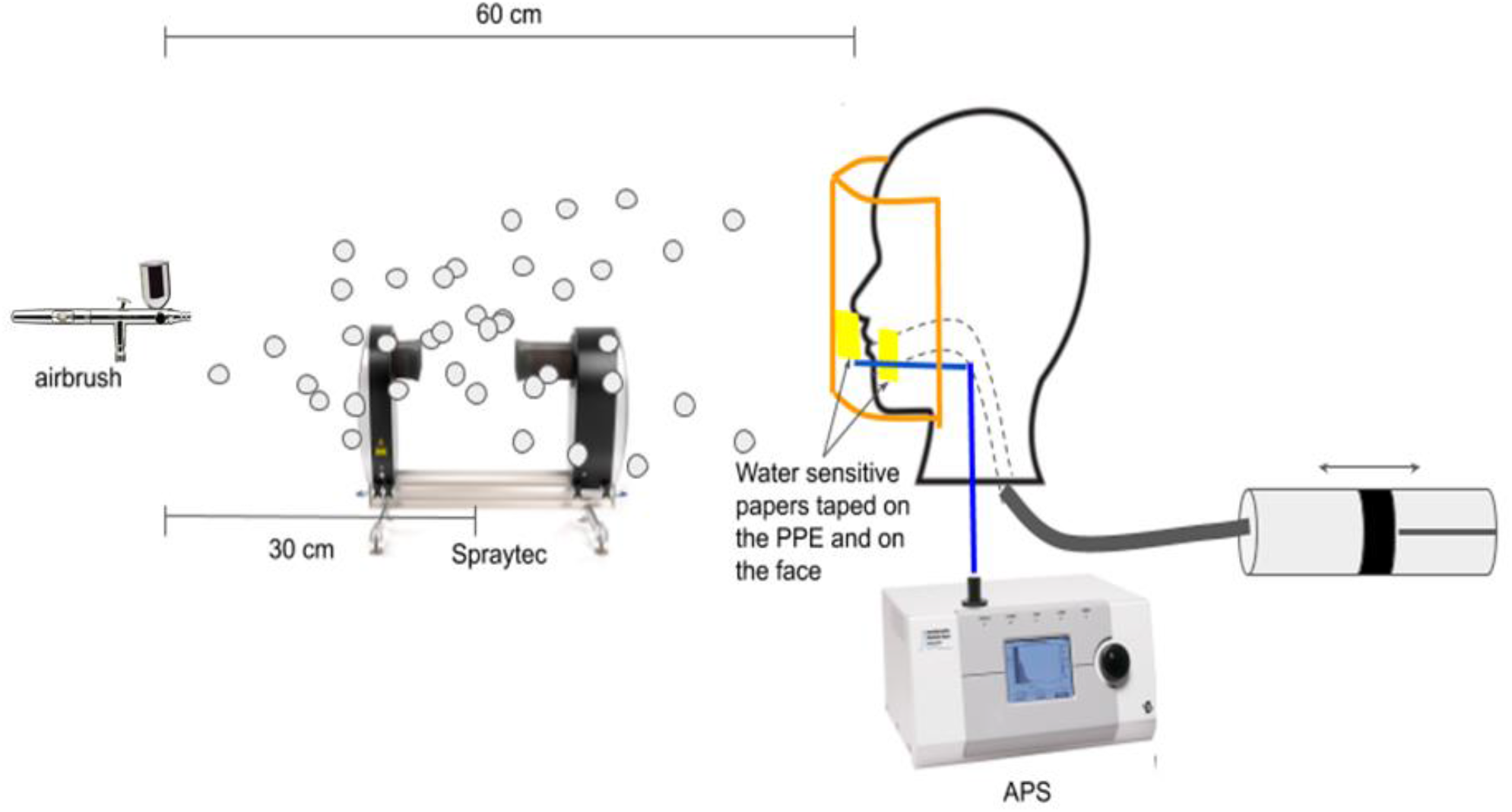
A sketch of the experimental setup. The manikin head is fitted with the PPE (in this case, a shield) and is connected to an oscillatory pump simulating breathing. The APS probe monitors the air very close to the mouth. The airbrush is located 60 cm away from the manikin head, and the ejected spray is monitored by the Spraytec instrument 30 cm away. In an alternative setup (not shown), the breathing machine was disconnected and the base of the manikin head was directly connected to the APS instrument.

**Figure 2.**
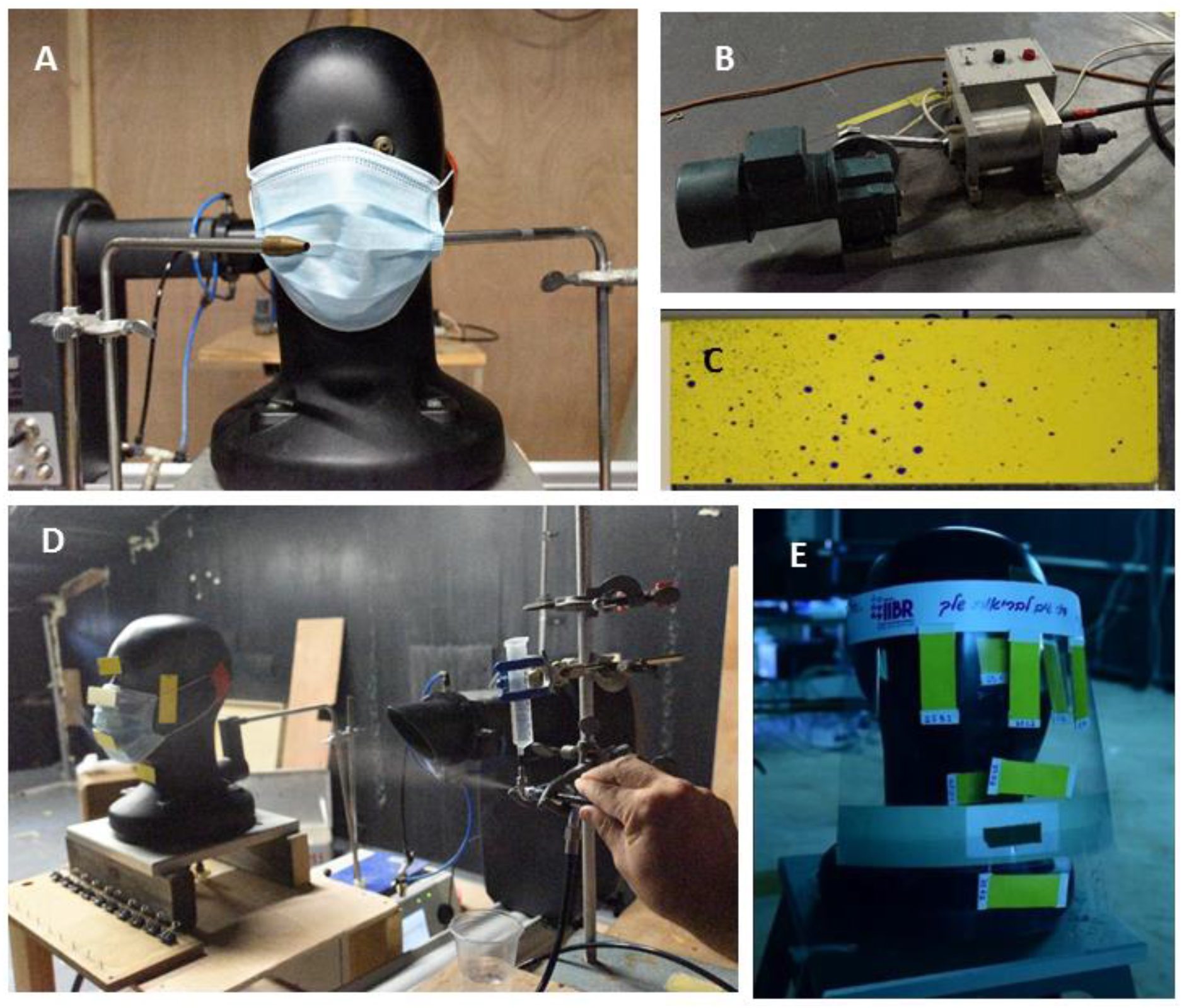
Elements of experimental setup. A) A manikin head with a fitted surgical mask. A probing tube for APS measurement is inserted through the right gap between the mask and the face. B) The breathing machine oscillatory pump. C) Water sensitive paper after exposure. D) A simulated expiratory event. The airbrush diffuser can be seen on the right. E) Water sensitive paper taped on the shield and on the manikin face underneath.

Adjusting the jet direction and estimating the spray exit velocity were done by high-speed photography using a Photron high-speed digital camera at 4500 frames/sec. The jet velocity was estimated by measuring the time for the plume to propagate a given distance. The instantaneous size distribution of airborne droplets produced by the airbrush diffuser was monitored by a laser diffraction system, “Spraytec” [Malvern Panalytical – Malvern, Worcestershire, UK]. Mass and number distributions as measured 30 cm from the diffuser are shown in Figure 3, where the different colors indicate different repetitions. Minor differences between repetitions arise mostly as a result of the unsteady flow generated by the jet. The spray number distribution is characterized by diameters of 1.29, 1.76, and 6.51 μm for the 10^th^, 50^th^, and 90^th^ percentiles, respectively. Additionally, the droplet volume distribution diameter is characterized by 8.87, 22.25, and 280.64 μm for the 10th, 50^th^, and 90^th^ percentiles, respectively. Three modes can be observed in the volume distribution, at 2, 20, and 620 μm, with Full Width Half Maximum (FWHM) of 1.5, 35, and 630 μm, respectively. The main mode in the number distribution is consistent with the reported measured distribution for cough events (Morawska et al., 2009)

**Figure 3.**
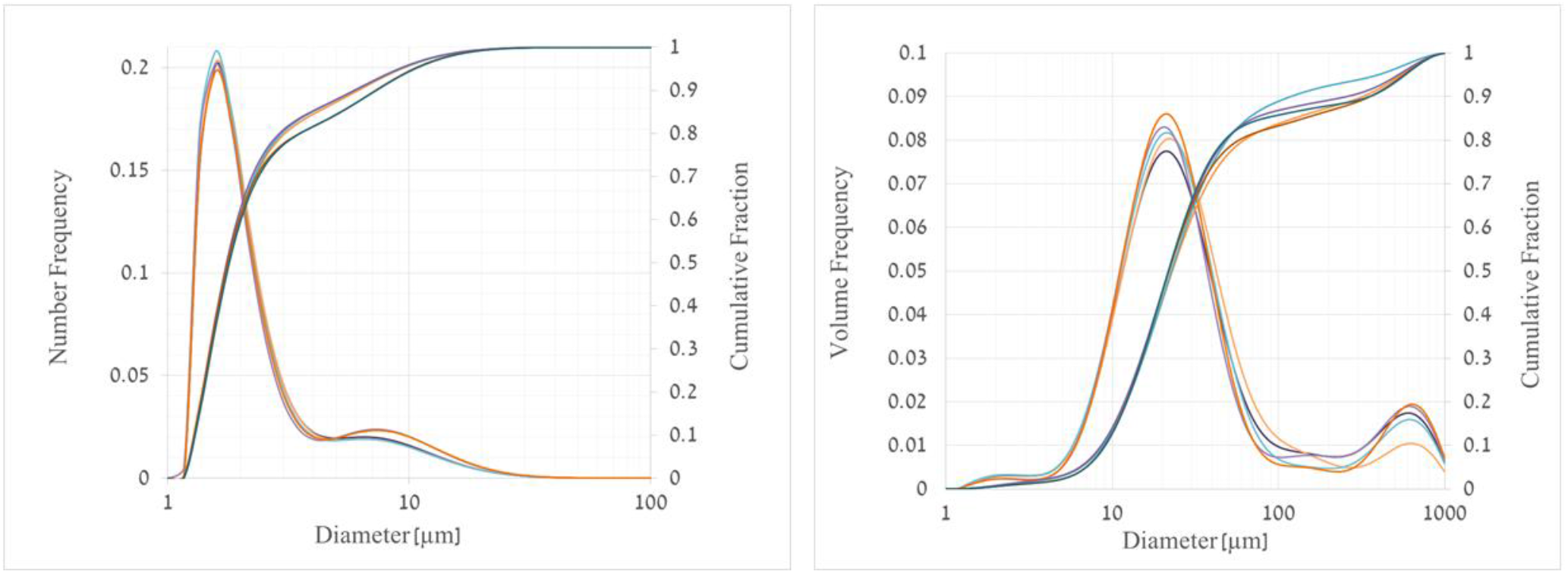
Spraytec measurements of number (left) and volume (right) size frequency and cumulative distributions of jet droplets, measured 30 cm from source. The different colors indicate different repetitions.

It is worth noting, that observed distributions of cough-generated droplets include sizes of up to a few hundred micrometers (Xie et al. 2009). In contrast, our cough simulator is limited to droplets smaller than 100 μm. However, large droplets are inertial and ballistic, and their increased presence in real cough events would therefore lead to an increase of the actual blocking efficiency.

A mechanical breathing machine (piston pump, self-production, see Figure 2), with a standard medium-sized manikin head [Dräger, Ltd.], located downstream from the diffuser, was used to simulate a respiring person. The breathing waveform was sinusoidal with a flow rate of 21 liters per minute (1.1 liter per a single breath). The simulated coughs were timed to the moment of maximal intake in the breathing simulator cycle.

The location of the breathing simulator was set at 60 cm, simulating a distance of mutually interacting individuals. In order to take into account height differences between the infected and the susceptible individuals, some trials were conducted where, besides the 60 cm horizontal distance, a vertical distance of 30 cm was set by shifting the cough simulator up or down.

The measurements were performed in two phases. The first phase was designed to assess the blocking efficiency against inhaled fine particles, while in the second phase, droplets potentially impacting the whole face were detected.

In the first phase, several tens of repetitions were conducted for each PPE, which included a surgical mask [Shengguan LTD.], two face shields (FS4 and FS5, see Table 1), and a N95 respirator (MOLDEX LTD.) which filters fine particles and was used as a reference. Additionally, some trials were conducted without any PPE.

**Table 1.** Sizes of the different plastic face shields used in the experiments. FS1 to FS4 are different commercial plastic face shields. FS5 is an extra wide plastic shield produced in-house for the trials.

| Face shield | Width (cm) | Height (cm) |
| --- | --- | --- |
| FS1 | 25.5 | 35.0 |
| FS2 | 32.0 | 25.0 |
| FS3 | 30.0 | 28.0 |
| FS4 | 28.0 | 28.0 |
| FS5 | 40.0 | 29.5 |

Tests were carried out mainly for a frontal configuration, in which the manikin head directly faced the airbrush. In order to test the significance of orientation, some tests were performed for a side configuration with the manikin head facing perpendicular to the jet direction. During the first phase, the breathing simulator and the cough simulator were at the same height.

Monitoring fine particle concentration was conducted using an APS 3321 instrument [TSI-Minneapolis, Minnesota, USA]. Two alternate setups were performed. In the first, the air very close to the mouth of the manikin head was sampled through a thin tube (see Figure 2). The tube was connected to the APS instrument by a 30 cm vertical segment, following a 20 cm horizontal segment extending to the exterior part of the mouth through the gap between the mask\shield and the head. In the second setup, the APS was directly connected to the base of the manikin head, instead of to the breathing machine. The APS maintains a steady flow rate of 5 l/min into the instrument.

For particles around 1 μm or larger, loss rate *β*(sec^-1^) of particles in the horizontal tube can be estimated (Lai and Nazaroff, 2000) using the approximation 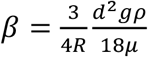, where *R* is the tube radii, *d* and *ρ* are the particle diameter and density, *μ* is the dynamic viscosity of air, and *g* the acceleration of gravity. The mean flow velocity inside the tube is around 1 m/s, thus the particles spend only about 0.2 seconds in the horizontal segment of the tube. Within that time, the fraction of particles deposited in the tube is *1* − exp(−*0*.*2β*), which, for particles of 2, 5, and 7 μm, yields tolerable loss fractions of 0.8%, 5%, and 9%, respectively.

The APS instrument monitors continuously, with a time resolution of 1 second, and provides the particle number concentration and size distribution (between 0.3 to 20 μm). To ensure sufficient concentration of very fine particles, the cough simulator used 0.25 gr/L NaCl in an aqueous solution.

In the second phase, yellow, water sensitive papers [Quantifoil] that change color upon contact with water provided an indication of droplets reaching different areas on the manikin head. The papers were taped (see Figure 2) on the shield and on the manikin face at six representative regions: forehead, nose, chin, left cheek, right cheek and neck. Different face shields were examined (Table 1), and again, for reference, a surgical mask was also tested. In the latter case, papers were taped on the nose and the chin. For some of the runs, the manikin head was shifted 30 cm up or down, or, alternatively, placed facing perpendicular to the jet. Each trial was repeated twice. After the absorption of droplets onto the water sensitive papers, the papers were photographed using a digital camera (Nikon D810) with macro lens (Nikon Macro D 105 mm). Following a former work (Lindsley, et al., 2014) which showed that about 90% of the total spray mass accumulates during the first few seconds, the papers were collected immediately after each trial, and photographed within one hour after exposure. The photographs were analyzed (by Image Pro Plus Version 10.0, Media Cybernetics, Maryland USA) to determine the number of droplets. In order to determine the smallest pre-impact diameter that could be identified on the water sensitive papers (in our case, 3 μm), we used the method described in Hoffmann and Hewitt (2005).

The protection efficiency of a face shield was assessed by testing its ability to block the droplets from reaching the face. This is achieved by counting the droplets on the face and on the shield for every specific run. Equation (1) defines the *blocking efficacy*, evaluated separately for each run, as the percentage of the number of droplets deposited on the shield (N_above_), relative to the total number of droplets on the shield and face (N_below_) together.

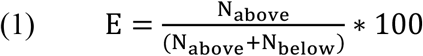

It is worth elaborating here on the utilization of Equation (1) in the current work. The straightforward way of evaluating the total mass of fluid impacting the face and shield is through addressing the size distribution of the droplets found on the papers. Here, an alternative and simpler way was followed: It is reasonable to assume that a small droplet, moving in the airstream, has a greater chance of reaching the face than a large inertial droplet flung at the shield. Therefore, the blocking efficacy, Equation (1), provides a lower bound for the percentage of mass of contaminated fluid blocked by the shield.

## RESULTS

### Fine particle penetration-phase 1

Figure 4 presents a typical APS signal showing 3 sequential repetitions. The typical time interval after a simulated cough event when a concentration above the room’s background could be identified is around one minute. Maximal concentrations were observed around 10 seconds after the cough event onset.

**Figure 4.**
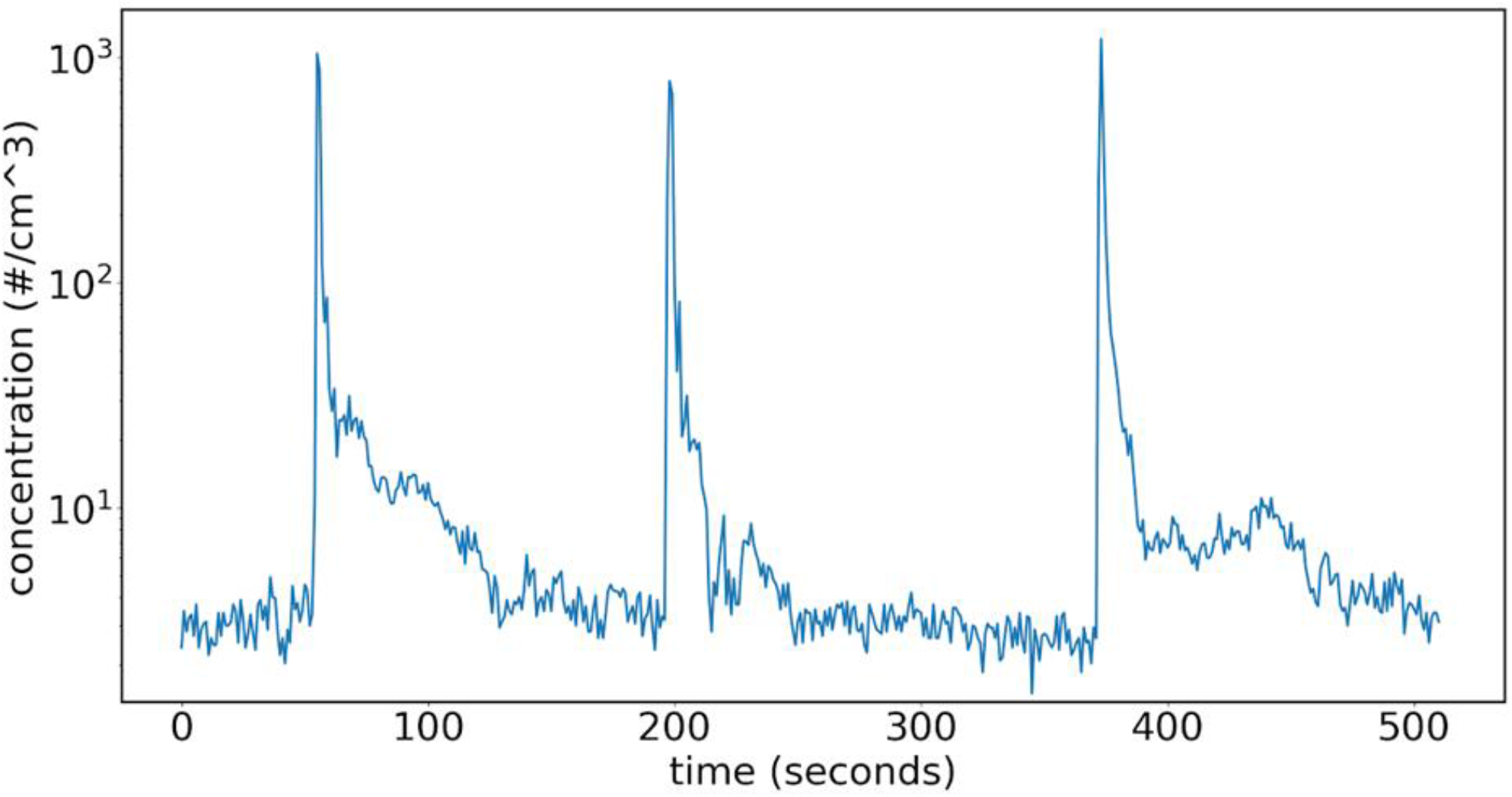
Example of typical APS number concentration signal. A few seconds after a cough is generated, the concentration sharply rises. Then, during the following minute it returns to the background values, around 2-3 particles cm^-3^.

The number of simulated cough events for each of the PPE tested during the first phase of measurement was 20, 21, and 11 for frontal exposure tests of the face shield, the surgical mask and the N95 respirator, respectively. Also, tests of a regular face shield (FS4), an extended face shield (FS5) and a surgical mask (5 tests each) were performed for side exposure. Additionally, 30 repetitions were conducted with no protection. Changes in the probing locations of the APS (see Methods) did not have a significant influence on the results and therefore the results are presented without addressing this issue.

As a simple measure of particle infiltration, we define *C*_*m*_(*n*), as the maximal number concentration for repetition *n* (peaks in Figure 4). In the same way, *C*_*m*_(*n, d*) is the maximal number concentration for a specific size bin, *d*. In order to compare different PPE or configurations, we also define an ensemble average which is the maximal number concentration for particles of size bin *d*, averaged over all *N*_*p*_ repetitions associated with a specific PPE or configuration (*p*).

Figure 5 shows 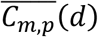 for a face shield as calculated by Equation (2), a surgical mask, an N95 respirator, and for the cases with no protection. For fine particles up to a diameter of about 2.2 μm (Figure 5A), the dependence of the concentration on particle size is qualitatively similar for the different cases, with a slight decrease in amplitude moving from no protection to surgical mask, and a more significant decrease moving further to the N95 respirator and the face shield tests. For particles larger than about 2.5 μm (Figure 5B), the surgical mask and the face shield cases exhibit similar behavior, with substantially lower concentrations, up to about two orders of magnitude, compared to the case of no protection. For these larger particles, the lowest concentration values were attained with the N95 respirator.

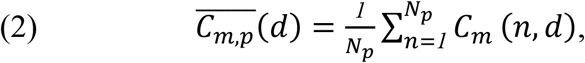

**Figure 5.**
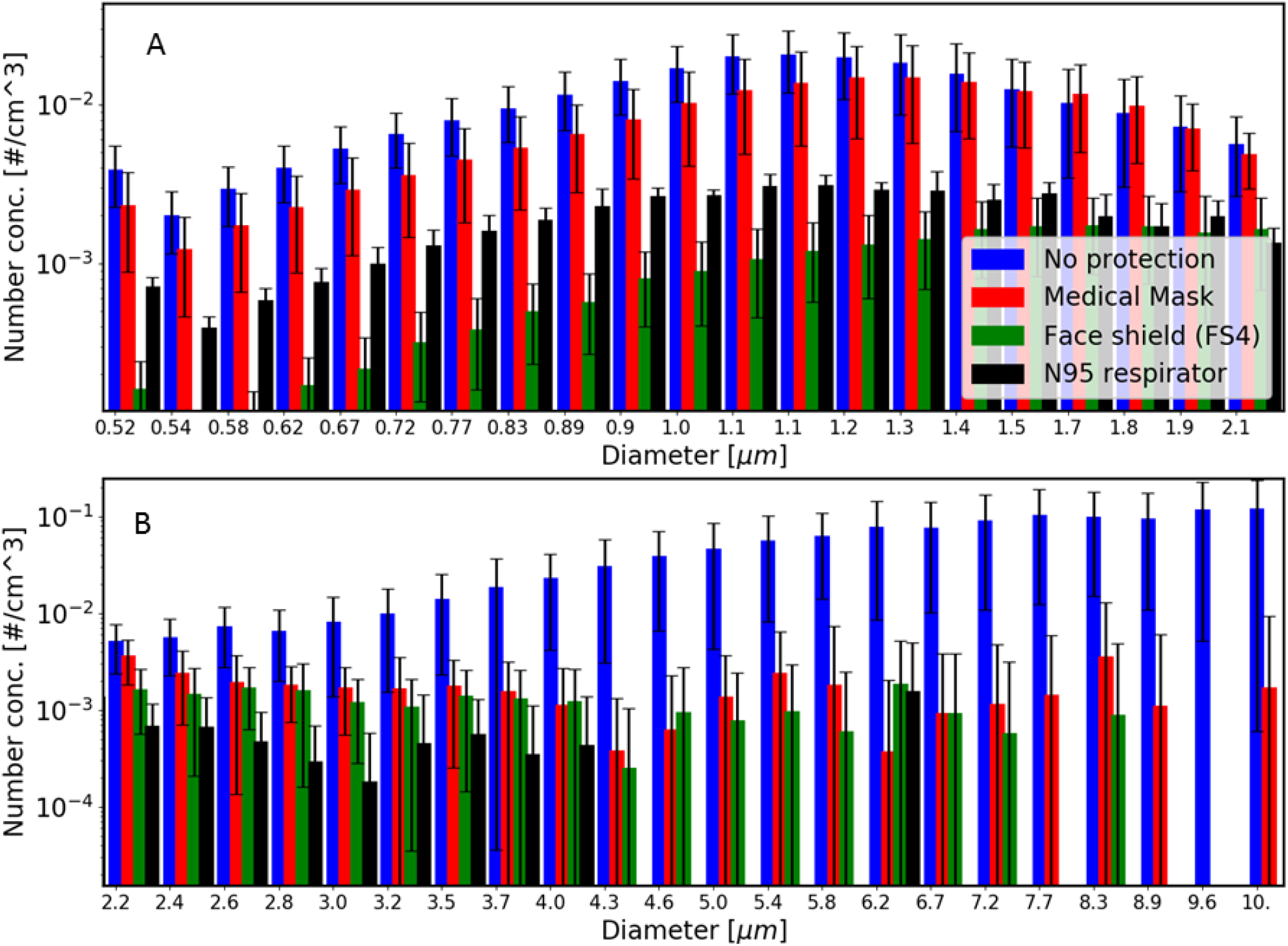
APS measurements statistics of inhaled particles number concentration, see Equation (2). The horizontal axis specifies the aerodynamic particle diameter (A-fine particles, B-large particles). Repetitions are grouped into four different cases as indicated by the different colors. The standard deviation among different repetitions in each group is indicated by error bars.

In Figure 6, *C*_*m*_ is shown for all repetitions (red circles). Horizontal red lines mark the median, the blue squares indicate the second and third quarter, and the extreme horizontal lines describe the first and ninth decile. Median values for cases with no protection, a shield, a surgical mask, and a N95 respirator are around 750, 40, 450, and 100 particles/cm^3^, respectively. Additionally, results for side exposures with a mask, a shield (FS4), and an extended shield (FS5) are shown. For these cases, median concentrations were 650, 2000, and 30 particles/cm^3^, respectively.

**Figure 6.**
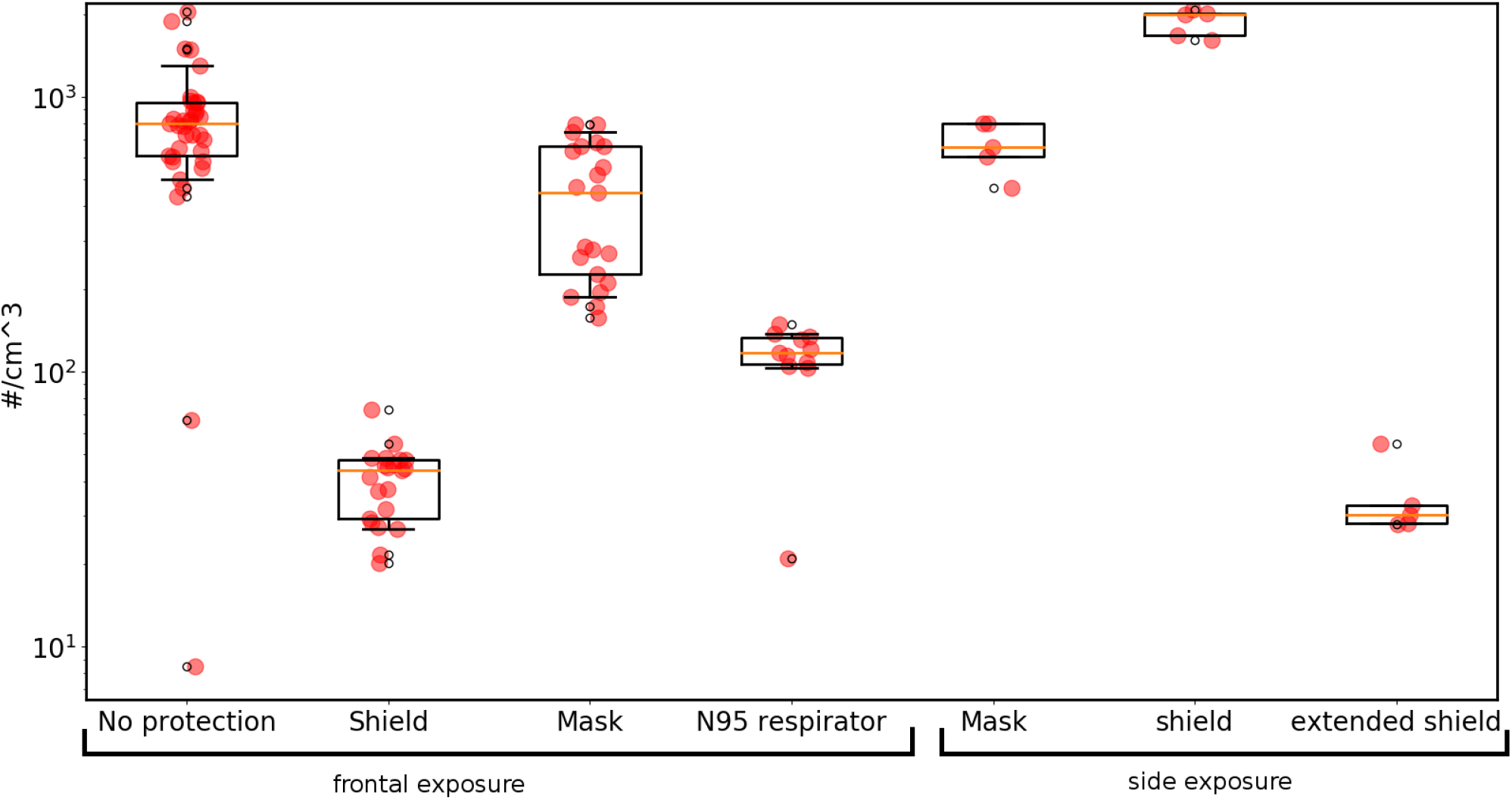
Peak number concentration statistics, as measured by the APS instrument. Each dot represents the peak concentration of one repetition. Repetitions are divided into frontal and side exposures; in the former the groups are in an un-protected scenario (32 repetitions), plastic shield (20 repetitions), surgical mask (21 repetitions), and a N95 respirator (11 repetitions), and in the latter, surgical mask, regular shield (FS4), and an extended shield (FS5), with 5 repetitions each. The horizontal red line marks the median, the blue square indicates the second and third quantiles, and the horizontal black lines show the first and ninth decile.

Despite the considerable scatter among different repetitions, a few conclusions are evident:

- For the frontal exposures, an advantage of face shields in comparison to surgical masks can be seen. While the surgical mask reduces the number of inhaled particles by roughly a factor of two, the face shield provides better protection and blocks more than 90% of the otherwise inhaled particles.
- The performance of the regular shield (FS4) in blocking particles dramatically depends upon the orientation of the manikin head. For side exposures, shield FS4 does not seem to provide any protection.
- The extended shield (FS5) provides good blocking even for side exposure.

### Droplet impaction-phase 2

During the second phase, the blocking efficacy was estimated through the analysis of water sensitive papers and applying equation (2). Figure 7 presents the overall average blocking efficacy for each tested scenario (excluding the neck area which was not covered completely).

**Figure 7.**
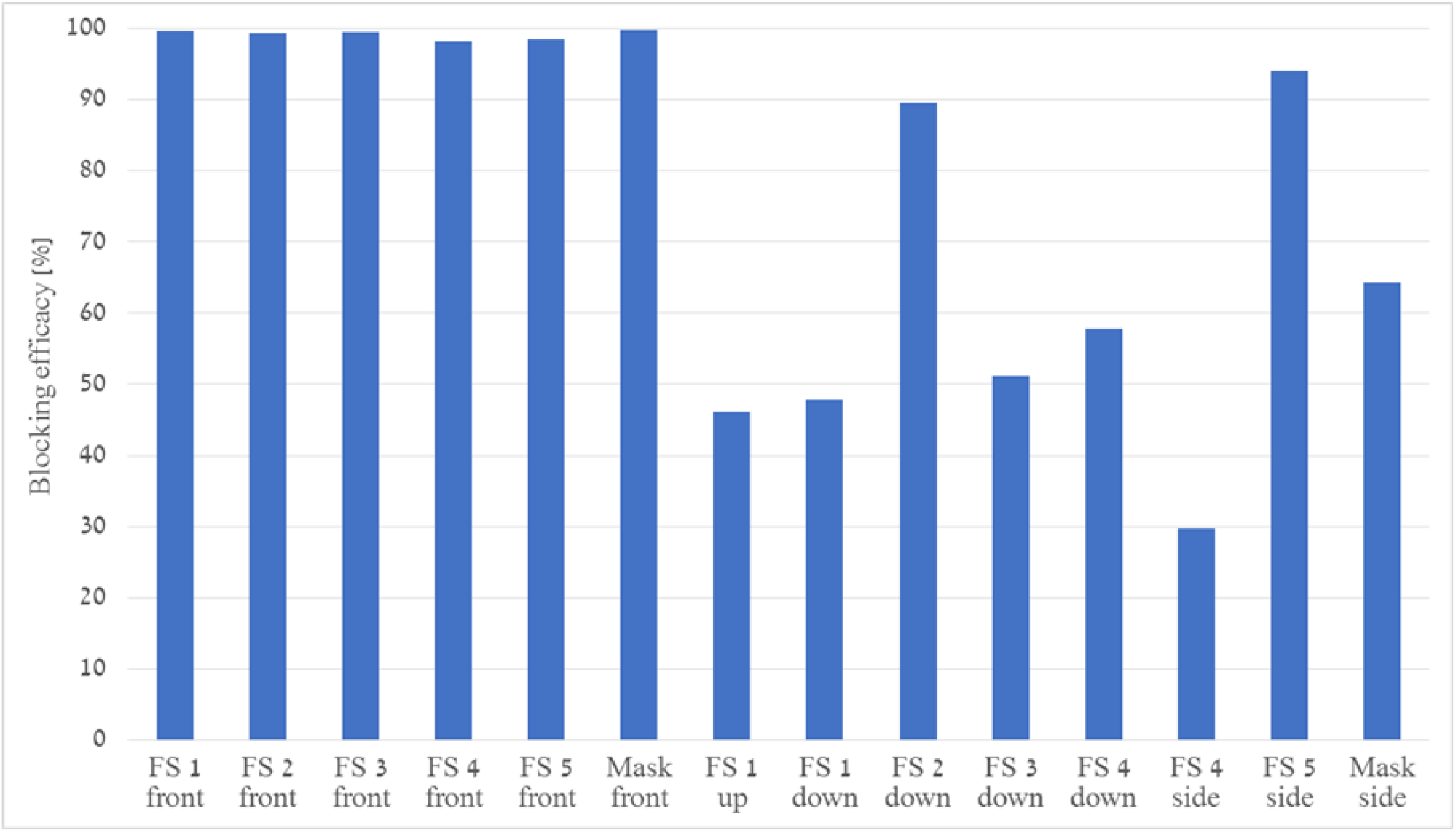
The blocking efficacy (equation 1) of different face shields (FS) and a mask, for different configurations. The labels indicate the specific configuration: ‘front’-frontal exposure, ‘up’(‘down’)-frontal configuration where the airbrush is moved 30 cm upward (downward). ‘side’-the manikin head faces perpendicular to the jet direction, with no vertical separation.

As evident from Figure 7, when frontal exposure is considered and without vertical separation, the blocking efficacy is higher than 90% for all face shields and also for the mask. As noted above, these findings apply to droplets of diameter larger than 3 μm, which is consistent with the APS observations (see Figure 5) showing similar blocking capabilities of the surgical mask and the face shield for large particles.

In contrast, in trials which include a vertical distance between the cough and breathing simulators, the blocking efficacy is as low as about 45%. However, one should note that for such events, the overall number of deposited droplets is relatively low.

The three bars on the far right in Figure 7 refer to side exposures. All PPE block fewer droplets in this orientation than in comparison with frontal exposure, possibly due to penetration of inertial droplets through the side gap (between the face and the mask/shield), which is directed towards the source. The potential advantage of properly designed visors is reflected by the improvement in the blocking efficacy of the extended shield (FS5), which is substantially higher than the corresponding value for a mask.

It is interesting to compare the efficacy for different facial areas. Figure 8 presents the blocking efficacy evaluated for specific areas. For cases without vertical separation, the efficacy depends on the location: For the center of the face, the forehead, the nose and the chin, it is close to 100%, while at the cheeks and neck, which are closer to the visor’s margins, the efficacy falls to 40-50%.

**Figure 8.**
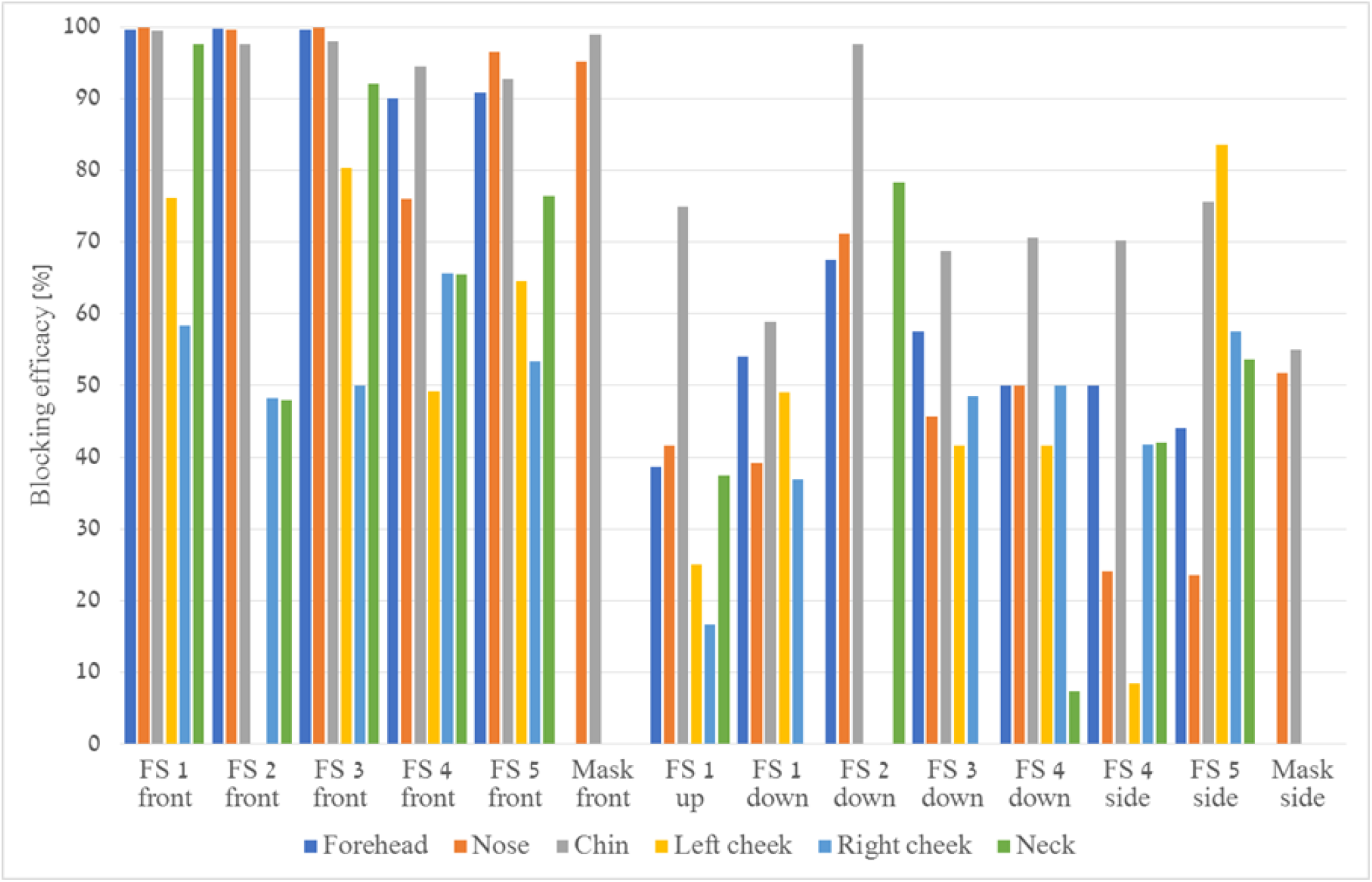
Blocking efficacy for specific facial areas and different PPE. The labels indicate the configuration: ‘front’-frontal exposure, ‘up’(‘down’)-frontal configuration where the airbrush is moved 30 cm upward (‘downward’). ‘side’-the manikin head faces perpendicular to the jet direction, with no vertical separation.

An added vertical distance of 30 cm above and below results in an increase in the relative penetration of droplets, regardless of the facial area. The droplets penetrate through the shield openings, and affect all parts of the face, not only those close to the openings.

For side orientation tests in which the left cheek faces the source (three groups of bars furthest to the right), a decrease in all efficacy values is observed. The enhanced blocking provided by the extended shield (FS5) is mostly manifested in the efficacies related to the left cheek (yellow bars), with an improvement from ∼10% to ∼85%.

### Protection provided to persons surrounding an infected wearer

The potential of a shield worn by an infected individual to prevent transmittance was tested by placing the shield on the diffuser in the location and orientation to simulate usage by an infected person. The manikin head was again positioned 60 cm away, facing the source. Water sensitive papers were taped on different manikin facial areas and APS measurements were taken. For comparison the same was repeated with a surgical mask. In both cases, 15 cough events were simulated sequentially, 30 seconds apart. After the series of coughs, the papers were analyzed. For both the shield and the mask cases, no indication of droplets could be found on the papers. Also, no signal higher than the background values could be detected by the APS instrument. This implies that for the configuration described, a shield worn by an infected individual seems to block the infected spray emitted in an expiratory event as effectively as a surgical mask.

## DISCUSSION

Motivated by the potential of mass-use of face shields in place of surgical masks to control the spread of COVID-19, various shields were tested. A carefully calibrated airbrush diffuser was used to simulate human coughs. A manikin head connected to a breathing simulator was used to examine different protective equipment, including a surgical mask, a N95 respirator, as well as different kinds of face shields. Different configurations and orientations were also tested. The penetrating air was monitored with an APS instrument for fine particles, and relative infiltration was assessed by analyzing water-sensitive papers that were taped on the face and on the PPE

For frontal exposure, the droplet blocking efficacy of different shields was comparable to that of a surgical mask. Fine particles were blocked even more efficiently by a shield than by a mask. However, the protection provided by a shield was found to depend on the exposure orientation and on the configuration. When vertical distance between the source and the manikin head was introduced, or alternatively, when the side of the manikin head is directed towards the source, the performance, relative to a mask, is degraded. Tests with an extended visor have shown to greatly improve the protection provided by the shield for side orientations. This result implies that extending shields toward covering a wider part of the face, especially the neck and the cheeks, may improve their efficacy. This is consistent with the recommendations of the Centers for Diseases Control and Prevention (Workplace Safety & Health Topics, 2015).

Protection provided to the persons surrounding the wearer was tested placing a shield and a mask on the source of the spray, and monitoring the manikin face for droplets and particles. After multiple consecutive simulated cough events, no droplets could be found on the water sensitive papers. Also, no concentration higher than the background concentration could be detected.

It is reasonable to assume that the efficient blocking exhibited by the face shield for fine particles is a consequence of a combination of the visor’s impermeability together with the flow and mixing patterns formed by the interaction of the jet and the shield. When the jet impacts the shield, it is deflected to the sides, away from the airways. At the visor’s margins, the abrupt change in the boundary layer induces turbulent mixing which lowers the concentration. Ambient air flow carries the contaminated air further away, and the fraction of eventually infiltrating particles is low.

A few comments regarding the limitations of the study and the interpretation of the results should be made:

First, the distribution of pathogens among different droplet size populations is yet unknown for the novel coronavirus, as is the minimal infective dose. Therefore, these factors are not taken into account in our analysis. The relation between blocking efficacy and transmittance probability is an important issue not addressed in this study.

Second, during their flight, droplets evaporate, and their diameter is reduced. This process is affected by the flow velocity, temperature, and relative humidity in the close proximity of the droplet, which can deviate from environmental background values due to the local impact of the droplet cloud itself (Bourouiba. 2020). Studying this effect requires careful monitoring and control on the environmental parameters along the droplets path. This, too, was not in the scope of the current work.

Another important comment regards the protection a face shield, or a surgical mask against air-suspended, fine aerosol. A plastic visor is not a filter, and fine contaminated particulate matter, carried with the airflow, can easily go around the face shield. Likewise, a surgical mask, though made of permeable material that can, by itself, act as a filter, is not designed to block fine aerosol from being inhaled. A recent study (Konda, et al., 2020) reports that even a minor gap, allowing the flow to bypass the filter, drastically decreases the filtering performance. This is consistent with the low Simulated Workplace Protection Factor (SWPF) of surgical masks compared to N95 respirators (Lawrence, et al., 2006). This implies that practically, both because of inherent fitting problems and because many users do not wear them in an optimal way, surgical masks hardly provide any protection against a fine aerosol. The advantage of face shields found in the current study is limited to the case of direct exposure to an expiratory event (in our case, a cough), where directional flow is involved in the process. For these events, the ability of a face shield to deflect the flow and to create mixing is the origin of advantageous performance over surgical masks.

In this respect, it is also important to emphasize that N95 respirators that are designed to fit the face and filter particulate matter, provide much better protection against suspended aerosol than both surgical masks and face shields.

## CONCLUSIONS

The applied generic setup and methodology can be used to test infiltration of particulate matter for different PPE in various configurations and scenarios. Overall, our results imply that a jet of particles formed by an expiratory event directed towards a shield is efficiently blocked, which results in protection comparable to that attained by a surgical mask, and even higher for fine particles. Ensuring optimal performance may involve design of shields with a wider visor. Also, a shield worn by an infected person seems to be at least as effective as a mask in preventing transmittance to nearby, unprotected individuals.

## Data Availability

Data is be available upon request

## ACKNOWLEDGEMENTS

This work was supported by the Israeli Ministry of Defense. The authors would like to thank Dr. Boaz Lev from the Israeli Ministry of Health for initiating the project and Dr. Yehuda Alexander for his review and helpful comments.

## REFERENCES

Centers for Disease Control and Prevention, and The National Institute for Occupational Safety and Health. 2015. Workplace Safety & Health Topics. Eye Protection for Infection Control. Accessed July 152015. http://www.cdc.gov/niosh/topics/eye/eye-infectious.html.

Bourouiba, Lydia. 2020. Turbulent Gas Clouds and Respiratory Pathogen Emissions: Potential Implications for Reducing Transmission of COVID-19. JAMA 323 (18): 1837–38. https://doi.org/10.1001/jama.2020.4756.

Fennelly, Kevin P. 2020. Particle sizes of infectious aerosols: implications for infection control. The Lancet Respiratory Medicine.

Gralton, J., Tovey, E., McLaws, M. L., & Rawlinson, W. D. 2011. The role of particle size in aerosolised pathogen transmission: a review. Journal of Infection, 62(1), 1–13.

Hoffmann, Wesley Clint, and Andrew J. Hewitt. 2005. Comparison of Three Imaging Systems for Water-Sensitive Papers. Applied Engineering in Agriculture 21 (6): 961–64.

Jones, Rachael M. Relative contributions of transmission routes for COVID-19 among healthcare personnel providing patient care. Journal of Occupational and Environmental Hygiene (2020): 1–8.

Konda, A., Prakash, A., Moss, G. A., Schmoldt, M., Grant, G. D., & Guha, S. (2020). Aerosol filtration efficiency of common fabrics used in respiratory cloth masks. ACS nano, 14(5), 6339–6347.

Lindsley, William G., Francoise M. Blachere, Robert E. Thewlis, Abhishek Vishnu, Kristina A. Davis, Gang Cao, Jan E. Palmer, et al. 2010. Measurements of Airborne Influenza Virus in Aerosol Particles from Human Coughs. PLoS ONE 5 (11). https://doi.org/10.1371/journal.pone.0015100.

Lai, Alvin CK, and William W. Nazaroff. “Modeling indoor particle deposition from turbulent flow onto smooth surfaces.” Journal of aerosol science 31.4 (2000): 463–476.

Lawrence, R. B., Duling, M. G., Calvert, C. A., & Coffey, C. C. (2006). Comparison of performance of three different types of respiratory protection devices. Journal of occupational and environmental hygiene, 3(9), 465–474.

Lindsley, William G., John D. Noti, Francoise M. Blachere, Jonathan V. Szalajda, and Donald H. Beezhold. 2014. Efficacy of Face Shields against Cough Aerosol Droplets from a Cough Simulator. Journal of Occupational and Environmental Hygiene 11 (8): 509–18. https://doi.org/10.1080/15459624.2013.877591.

Lindsley, William G., Jeffrey S. Reynolds, Jonathan V. Szalajda, John D. Noti, and Donald H. Beezhold. 2013. A Cough Aerosol Simulator for the Study of Disease Transmission by Human Cough-Generated Aerosols. Aerosol Science and Technology 47 (8): 937–44. https://doi.org/10.1080/02786826.2013.803019.

Morawska, Lidia, and Donald K. Milton. 2020. It is time to address airborne transmission of COVID-19. Clin Infect Dis 6: ciaa939.

Morawska, L., G. R. Johnson, Z. D. Ristovski, M. Hargreaves, K. Mengersen, S. Corbett, C. Y.H. Chao, Y. Li, and D. Katoshevski. 2009. Size Distribution and Sites of Origin of Droplets Expelled from the Human Respiratory Tract during Expiratory Activities. Journal of Aerosol Science 40 (3): 256–69. https://doi.org/10.1016/j.jaerosci.2008.11.002.

Perencevich, Eli N, Daniel J Diekema, and Michael B Edmond. 2020. Moving Personal Protective Equipment Into the Community: Face Shields and Containment of COVID-19. JAMA 323 (22): 2252–53. https://doi.org/10.1001/jama.2020.7477.

Roberge, Raymond J. 2016. Face Shields for Infection Control: A Review. Journal of Occupational and Environmental Hygiene 13 (4): 239–46. https://doi.org/10.1080/15459624.2015.1095302.

Tang, Julian W., Andre D. Nicolle, Christian A. Klettner, Jovan Pantelic, Liangde Wang, Amin Bin Suhaimi, Ashlynn Y.L. Tan, et al. 2013. Airflow Dynamics of Human Jets: Sneezing and Breathing - Potential Sources of Infectious Aerosols. PLoS ONE 8 (4): 1–7. https://doi.org/10.1371/journal.pone.0059970.

Xie, Xiaojian, Yuguo Li, Hequan Sun, and Li Liu. 2009. Exhaled Droplets Due to Talking and Coughing. Journal of the Royal Society Interface 6 (SUPPL. 6). https://doi.org/10.1098/rsif.2009.0388.focus.

Yang, Shinhao, Grace W.M. Lee, Cheng Min Chen, Chih Cheng Wu, and Kuo Pin Yu. 2007. The Size and Concentration of Droplets Generated by Coughing in Human Subjects. Journal of Aerosol Medicine: Deposition, Clearance, and Effects in the Lung 20 (4): 484–94. https://doi.org/10.1089/jam.2007.0610.

